# Correlates of Risk for Disinhibited Behaviors in the Million Veteran Program Cohort

**DOI:** 10.1101/2023.03.22.23286865

**Authors:** Peter B. Barr, Tim B. Bigdeli, Jacquelyn L. Meyers, Roseann E. Peterson, Sandra Sanchez-Roige, Travis T. Mallard, Danielle M. Dick, K. Paige Harden, Anna Wilkinson, David P. Graham, David A. Nielsen, Alan Swann, Rachele K. Lipsky, Thomas Kosten, Mihaela Aslan, Philip D. Harvey, Nathan A. Kimbrel, Jean C. Beckham the Million Veteran Program (MVP) and Cooperative Studies Program (CSP) #572

## Abstract

**Background:** Many psychiatric outcomes are thought to share a common etiological pathway reflecting behavioral disinhibition, generally referred to as externalizing disorders (EXT). Recent genome-wide association studies (GWAS) have demonstrated the overlap between EXT and important aspects of veterans’ health, such as suicide-related behaviors, substance use disorders, and other medical conditions.

**Methods:** We conducted a series of phenome-wide association studies (PheWAS) of polygenic scores (PGS) for EXT, and comorbid psychiatric problems (depression, schizophrenia, and suicide attempt) in an ancestrally diverse cohort of U.S. veterans (*N* = 560,824), using diagnostic codes from electronic health records. We conducted ancestry-specific PheWASs of EXT PGS in the European, African, and Hispanic/Latin American ancestries. To determine if associations were driven by risk for other comorbid problems, we performed a conditional PheWAS, covarying for comorbid psychiatric problems (European ancestries only). Lastly, to adjust for unmeasured confounders we performed a within-family analysis of significant associations from the main PheWAS in full-siblings (N = 12,127, European ancestries only).

**Results:** The EXT PGS was associated with 619 outcomes across all bodily systems, of which, 188 were independent of risk for comorbid problems of PGS. Effect sizes ranged from OR = 1.02 (95% CI = 1.01, 1.03) for overweight/obesity to OR = 1.44 (95% CI = 1.42, 1.47) for viral hepatitis C. Of the significant outcomes 73 (11.9%) and 26 (4.5%) were significant in the African and Hispanic/Latin American results, respectively. Within-family analyses uncovered robust associations between EXT and consequences of substance use disorders, including liver disease, chronic airway obstruction, and viral hepatitis C.

**Conclusion:** Our results demonstrate a shared polygenic basis of EXT across populations of diverse ancestries and independent of risk for other psychiatric problems. The strongest associations with EXT were for diagnoses related to substance use disorders and their sequelae. Overall, we highlight the potential negative consequences of EXT for health and functioning in the US veteran population.

## INTRODUCTION

Psychiatric disorders have far-reaching consequences for affected individuals, their families, communities, and the broader society ^1–4^. Many of these disorders are strongly co-morbid and share, at least in part, a common etiology ^5^. Disorders related to behavioral disinhibition, such as substance use disorders (SUD), conduct disorder, and antisocial personality disorder, have been labeled as externalizing disorders ^6,7^. Twin and family studies suggest that the common liability towards externalizing disorders is highly heritable (∼80%) ^8–10^. Recent multivariate genome wide association studies (GWAS) have found robust evidence for a latent genomic factor for externalizing disorders ^11,12^, composed of input GWAS related to substance use disorders, risky sexual behaviors, personality characteristics, and neurodevelopmental disorders. Importantly, genetic liability for externalizing disorders overlaps with other phenotypes of public health relevance, such as suicidal thoughts and behaviors ^11,13–15^, SUDs ^11,16–18^, and a range of other medical conditions (e.g., ischemic heart disease, liver disease, viral hepatitis) ^11^. The widespread impact of risk for externalizing disorders makes it a potential target for early intervention and prevention.

The proliferation of large-scale biobanks – such as All of Us ^19^, the UK Biobank ^20^, FinnGen ^21^, and Biobank Japan ^22^ – linking individual-level genomic data with electronic health records (EHRs) presents opportunities to further explore the relationships between genetic liability for a given disorder (typically in the form of polygenic scores, or PGS), and a wide range of clinical phenotypes. This hypothesis free approach, referred to as a phenome-wide association study (PheWAS) ^23^, can aid in identifying novel trait associations and understanding pleiotropic effects. Recent PheWAS using PGS for other psychiatric problems (e.g., schizophrenia, bipolar disorder, and depression) have identified widespread associations between PGS and a host of psychiatric and other medical diagnoses ^24,25^.

In the current analysis, we applied a PheWAS of a PGS derived from a multivariate GWAS of externalizing disorders/problems (EXT) ^11^ to the EHRs of the Department of Veterans Affairs Million Veterans Program Cohort (MVP) ^26^. A previous PheWAS of the EXT PGS in the Vanderbilt University Medical Center Biobank (BioVU) ^27^ identified over 250 associations with EHR derived medical conditions ^11^, but was limited to individuals of European ancestries. We extend the PheWAS of the EXT PGS all veterans of European, African, and Hispanic/Latin American ancestries. We further compared results from our primary PheWAS of EXT to analogously derived results for PGSs of schizophrenia (SCZ) ^28^, depression (DEP) ^29^, and suicide attempt ^13^, including joint modeling of these PGS. Finally, we attempted to replicate the findings from the primary PheWAS in a holdout sample of related veterans (full siblings). Genetic differences between siblings are random; therefore, within sibship associations between the EXT PGS and health outcomes cannot be attributed to between-family sources of confounding, such as environmental exposures that are correlated with population stratification.

## METHODS

### The Million Veterans Program Cohort (MVP)

Launched in 2010, MVP is a landmark endeavor that links genomic laboratory testing, survey-based self-report data, and EHRs, with the goal of creating a “mega-biobank” and evidence base for precision medicine initiatives ^26^. The 850,000 enrolled participants reflect the population that utilizes the Veterans Health Administration (VHA), with over-representation of older and male individuals, as well as higher rates of multiple morbidities and chronic conditions related to externalizing compared to the general population ^30,31^. Participants are active users of the VHA healthcare system and were recruited through invitational mailings or by MVP staff while receiving clinical care. Informed consent and authorization per the Health Insurance Portability and Accountability Act (HIPPA) were the only other inclusion criteria. Once enrolled, participants EHR data are linked, including diagnostic codes, routine laboratory results, and medications. The current analysis uses Release 4 of MVP data, as collection is ongoing. The present analyses were approved by the VA Central Institutional Review Board (IRB), and all participants provided written informed consent.

### Genotyping

MVP participants were genotyped on the MVP 1.0 Axiom array ^32^. Genetic ancestries of participants were classified using the HARE (harmonized ancestry and race-ethnicity) method ^33^, which harmonizes the closest ancestral population with self-identified race and ethnicity. Genotypic data were imputed to the Trans-Omics for Precision Medicine (TOPMed) reference panel ^34^, which specifically improves imputation quality in non-European and admixed ancestries ^35^. As of Release 4, there are 467,101 veterans of predominantly European ancestries (EUR), 124,717 veterans of predominantly African ancestries (AFR), 52,416 veterans of predominantly Hispanic/Latin American ancestries (HIS), and 8,362 veterans of predominantly Asian ancestries (ASN) with available genotypic and electronic health record data. In the current analysis, we included data from the EUR, AFR, and HIS groups, as these had sufficient statistical power for the number of associations tested. Within each of these HARE categories, we restricted analyses to unrelated individuals, excluding all those who were second-degree relatives or closer (KING coefficient ≤ 0.177) and limited to those whose primary self-identified race-ethnicity matched their HARE classification, so as to not introduce potential confounding driven by well-characterized health disparities related to racism and racial discrimination ^36,37^.

### Electronic health records (EHRs)

Our main outcomes for the analyses were phecodes, which are clusters of ICD-9/10-CM codes in the EHR ^38,39^ and have been validated previously ^40,41^. We considered individuals as having a diagnosis for any given phecode if there were two or more occurrences of that phecode in their EHR. Prior analyses have shown that 2 or more phecodes as a good predictor diagnosis ^24,25^. Phecodes are grouped into 17 categories: infectious diseases, neoplasms, endocrine/metabolic, hematopoietic, mental disorders, neurological, sense organs, circulatory system, respiratory, digestive, genitourinary, pregnancy complications, dermatologic, musculoskeletal, congenital anomalies, symptoms, and injuries & poisonings. We excluded phecodes for which there were fewer than 100 cases with the diagnosis. In total, there were 1,652 phecodes, 1,436 phecodes, and 1,125 phecodes with *N* ≥ 100 diagnoses available for EUR, AFR, and HIS veterans, respectively.

### Polygenic scores (PGS)

We estimated PGSs derived from four large-scale GWASs: externalizing problems (EXT, *N*_effective_= 1,492,085) ^11^, depression (DEP, *N*_effective_ = 449,856) ^29^, schizophrenia (SCZ, *N*_effective_ = 117,498) ^28^, and suicide attempt (SUI, *N*_effective_ = 91,230) ^13^. We focus on these additional PGS because: 1) internalizing (e.g., depression) and thought (e.g., schizophrenia) disorders are genetically correlated with EXT ^11^ and could confound associations between EXT and phenotypes of interest. 2) We included the SUI PGS, specifically, to explore the possibility of suicide risk related to impulsivity and behavioral disinhibition, independent of risk for specific to SUI or related to DEP. While PheWAS is an agnostic approach, included the additional measures of genetic liability to explore the robustness of potential associations with phenotypes of particular public health relevance to the veteran population (e.g., suicidal behaviors).

In EUR ancestries, we created PGS using PRS-CS ^42^, a Bayesian regression and continuous shrinkage method that uses an external reference panel (e.g., 1000 Genomes Phase III European subsample) to estimate the posterior effect sizes for each SNP in a given set of GWAS summary statistics. In the AFR and HIS ancestries, we used a different approach. PGS accuracy decays continuously as target samples differ in ancestry from the discovery GWAS, even within relatively homogenous genetic clusters ^43^ and we lacked ancestry-matched GWAS for EXT to use methods that boost power of PGS in underpowered samples, such as PRS-CSx ^44^. Therefore, in the AFR and HIS ancestries, we created EXT PGS using the 579 loci reported in the externalizing GWAS, as using genome wide significant variants is more robust to population stratification ^45^. We standardized all PGS to Z-scores.

### Analytic plan

We first conducted a phenome-wide association study (PheWAS) to examine the association between the EXT PGS and 1,652 phecodes within EUR ancestries, using logistic regression and covarying for age, sex, and twenty ancestry principal components (PCs). We also conducted the same PheWAS for the DEP, SCZ, and SUI PGSs to compare the overlap of significant associations from the four independent PheWAS. Next, we attempted to replicate any phenome-wide significant associations within AFR and HIS ancestries. Third, we performed a conditional PheWAS, including all PGS in the same model, to test whether associations between EXT remained while controlling for DEP, SCZ, and SUI PGSs. We also co-varied for total comorbidity burden, created by tabulating the number of unique phecode terms for which an individual met criteria for the 600 top parent codes, and then transforming this count using an inverse normal transformation ^25^. Lastly, to control for additional confounders, we used a subset of full siblings (*N* = 12,127) identified through genetic data, to test for associations between EXT and phecodes within-family. For within-family models, we used a linear probability model as opposed to logistic regression, as the fixed effects logistic regression can provide biased estimates when the number of observations per group is small ^46^. We applied a multiple testing correction for 1,652 tests across the 4 PGS included in the analyses for EUR ancestries (*p* < .05/6608 = *p* < 7.57×10^-4^). We used a less conservative approach to multiple testing in the AFR and HIS ancestries given the expected reduction in predictive power of PGS, applying a false discovery rate (FDR) ^47^ of 5%.

## RESULTS

### Main PheWAS of EXT PGS

Of the 467,101 veterans of broadly European ancestries (EUR), we filtered down to a sample of 438,384 unrelated individuals. After removing those with any missing information, those without available EHR data, and retaining those whose primary identity was Non-Hispanic White, *N* = 406,254 participants were available for the initial PheWAS in EUR (mean age = 69.8, SD = 14.1; 92.8% male). We performed a similar process for veterans of African (AFR, *N* = 112,390, mean age = 63.5, SD = 12.6; 86.3% male) and Hispanic/Latin American (HIS, *N* = 42,179, mean age = 60.5, SD = 16.2; 90.4% male) ancestries. eTable 1 presents all demographic statistics.

Of the 1,652 total phecodes we tested, 619 (37.5%) were significantly associated with EXT PGS after correcting for multiple testing. We observed significant associations for EXT PGS across all virtually all bodily systems, with the strongest positive associations being between EXT PGS and *viral hepatitis C* (OR = 1.49; 95% CI = 1.46, 1.51), *substance addiction and disorders* (OR = 1.39; 95% CI = 1.37, 1.40), *embolism/thrombosis of the abdominal aorta* (OR = 1.36; 95% CI = 1.22, 1.51), *tobacco use disorders* (OR = 1.35; 95% CI = 1.34, 1.36), and *cancer of the mouth* (OR = 1.33; 95% CI = 1.17, 1.50). We also observed robust negative associations with *autism* (OR = 0.76; 95% CI = 0.68, 0.86), *intestinal infection* (OR = 0.85; 95% CI = 0.80, 0.90), *disorders of bilirubin excretion* (OR = 0.90; 95% CI = 0.86. 0.94), and *prostate cancer* (OR = 0.97; 95% CI = 0.95, 0.98).

Figure 1, Panel A presents the distribution in EXT effect sizes (odds ratios, ORs) for all 619 significant PheWAS associations by phecode domain. Across domain, there is a general enrichment for positive associations with EXT: the median effect sizes are generally above one and greater levels of EXT risk are generally associated with increased risk of diagnoses. The one exception is pregnancy complications, which had no significant associations and is not unexpected given the predominantly male composition of this study sample. While there was variation in effect sizes across groupings of phecodes, the largest associations, on average, were for phecodes related to respiratory issues, mental disorders, injuries & poisonings, and infectious diseases (median ORs = 1.12 – 1.14).

**Figure 1:**
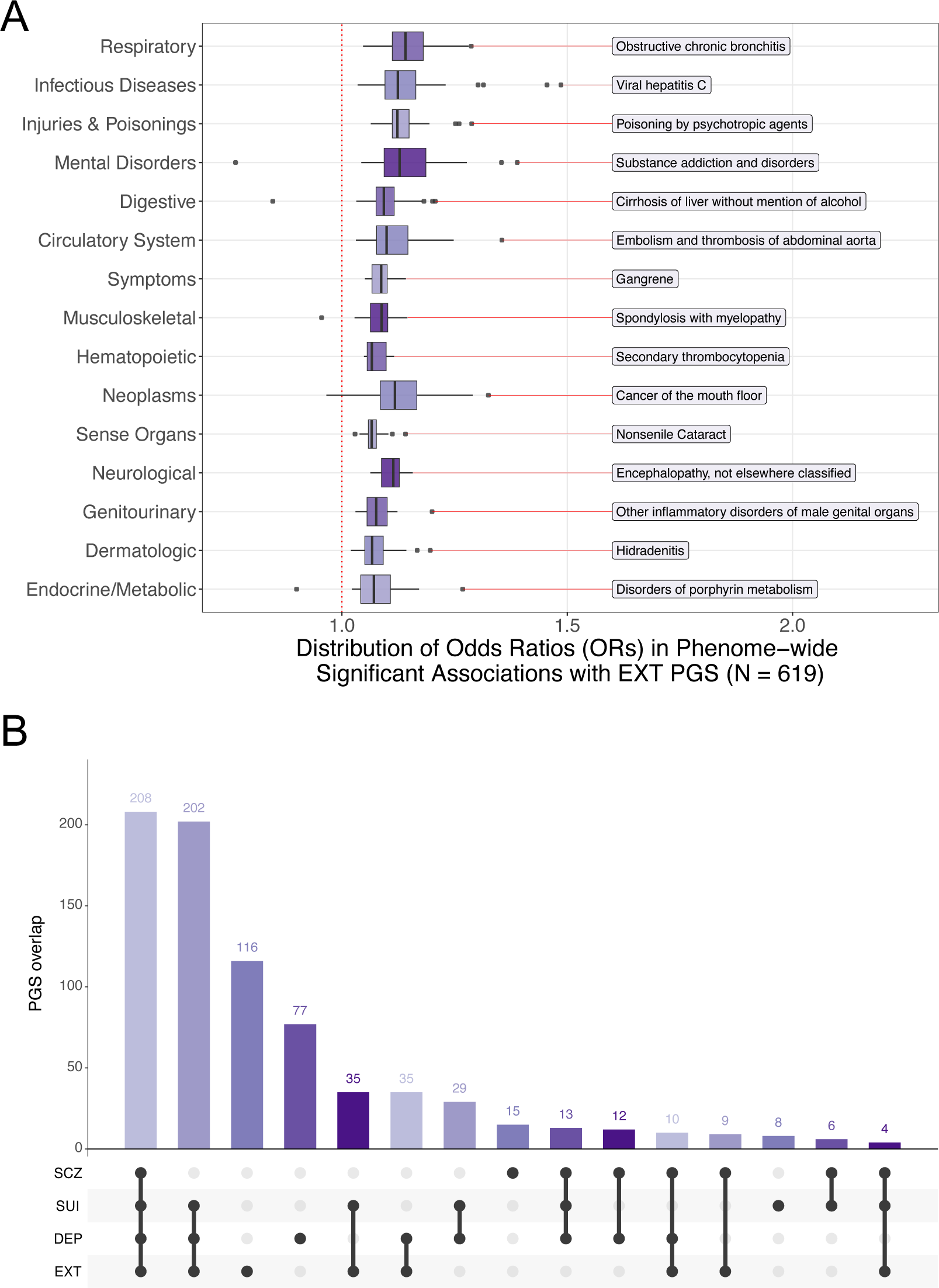
PheWAS of Externalizing Polygenic Risk in MVP. PheWAS associations with EXT PGS in veterans of the primarily European ancestries (N = 406, 254). Panel A presents the box plots of effect sizes (odds ratios, OR) for the 619 (out of 1,652) significant PheWAS associations below the Bonferroni corrected p-value threshold (*p* < 7.57*10^-6^). Panel B presents an upset plot of overlap between phenome wide significant associations (*p* < 7.57*10^-6^) across all four PGS (EXT, DEP, SCZ, and SUI).

Lastly Panel B presents the breakdown of independent phenome-wide significant associations across the EXT, DEP, SCZ and SUI PGS^48,49^. Across all the 6,608 tests (4 PGS with 1,652 phecodes), 779 (11.78%) of the associations had p-values below the Bonferroni corrected threshold (*p* < 7.58×10^-4^). The majority of these associations (56.1 %, N = 437) involved three of the four PGSs, and over 70% (N = 561) involved two or more of the PGSs. The full results are in eTable 2.

### Multi-Ancestry Verification of EXT PGS

We next examined whether EXT PGS associations that were significant in the main EUR analyses were comparable in the AFR and HIS MVP participants. Among the 619 significant associations in veterans of EUR ancestries, 614 were available (*N*_DX_ ≥100) and 73 (11.9%) were significant in veterans of AFR ancestries, 584 were available and 26 (4.5%) were significant in veterans of HIS ancestries after correcting for multiple testing. Panel A in Figure 2 presents the proportion of significant association identified in the EUR PheWAS that replicated in either AFR or HIS ancestries, by phecode domain. Of all the domains, phecodes related to *mental disorders* had the highest of proportion of associations that replicated across ancestry (∼25%). Panel B includes a subset of these replicated associations, which included SUDs (alcohol, tobacco, and other substances), viral hepatitis, and problems related to the respiratory system (e.g., cancer, chronic airway obstruction, and respiratory failure). As expected, while these associations were significant, there was a large attenuation in effect sizes, consistent with using PGS derived from ancestries that differ from the target sample ^44,45^ (full results in eTable 3).

**Figure 2:**
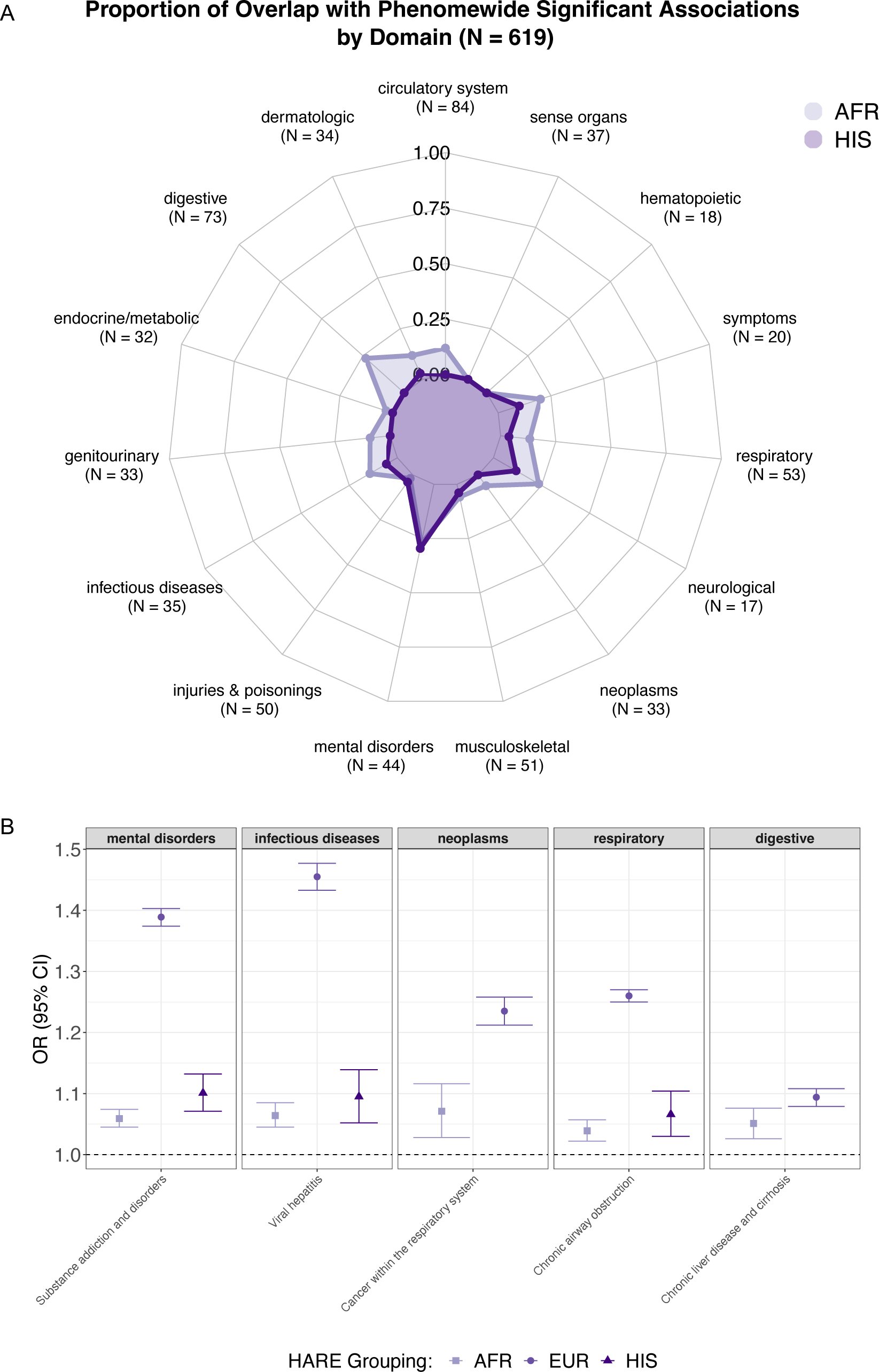
Multi-Ancestry Results for Externalizing Polygenic Risk in MVP. Overlap in associations across European (EUR), African (AFR), and Hispanic/Latin American (HIS) ancestries. Panel A present the proportion of PGS identified in EUR ancestries that were significant in the AFR and HIS groupings, by phecode domain. Numbers in parentheses represent the total number of significant associations in EUR, per phecode domain. Panel B presents selected associations and corresponding effect sizes (odds ratios, OR) of EXT PGS associations that replicated in either African or Hispanic/Latin American ancestry groups.

### Joint PheWAS of EXT, DEP, SCZ, and SUI PGSs

Next, we investigated whether the 619 phecodes associated with EXT PGS remained associated in models after conditioning on the DEP, SCZ, and SUI PGS. Of the 619 associations with EXT, 494 (79.8%) remained associated after conditioning on the other PGS and correcting for multiple testing (*p* < .05/619 = *p* < 8.08×10^-5^). Effect sizes ranged from 1.02 for overweight/obesity (phecode = 278; 95% CI = 1.01, 1.03) to 1.44 for viral hepatitis C (phecode 070.3; 95% CI = 1.42, 1.47) for traits positively related to the EXT PGS. Effect sizes for negative associations ranged from 0.74 for a diagnosis of autism (phecode = 313.3; 95% CI = 0.65, 0.84) to 0.97 for melanomas of the skin (phecode = 172.1; 95% CI = 0.96, 0.99). Associations that were no longer significant spanned all bodily systems, and included schizophrenia, rheumatoid arthritis, and chronic sinusitis, among many others. The median OR for the EXT PGS dropped from 1.10 in the marginal associations with these phecodes to 1.08 after conditioning on the other PGSs. Figure 2B presents a subset of the larger associations (OR >1.15, full results in Supplementary Table 4). While there was attenuation in effect sizes, EXT remained associated with the various phecodes independent of risk for DEP, SCZ, or SUI.

To examine whether EXT was associated with any of the 619 outcomes due to the increased number of comorbidities associated with higher levels of externalizing, we ran the PheWAS including a covariate for total comorbidity burden in veterans of European ancestries. Figure 3, Panel B includes box plots for the main PheWAS (EXT PGS only), the conditional PheWAS (EXT, DEP, SCZ, and SUI PGSs), and the model with all PGSs and the comorbidity score. There is an attenuation in median effect size across each set of analyses. Of the 619 significant associations identified in the mina PheWAS, 188 remained associated with EXT after adjusting for total comorbidity burden and the additional PGSs, with the top associations remaining those for viral hepatitis, substance use disorders, and complications from smoking (full results in eTable 5). Of these 188 scores, 181 (96%) were not phenotypes directly related to EXT (e.g., SUDs) and 165 (88%) were outside the domain of *mental disorders*.

**Figure 3:**
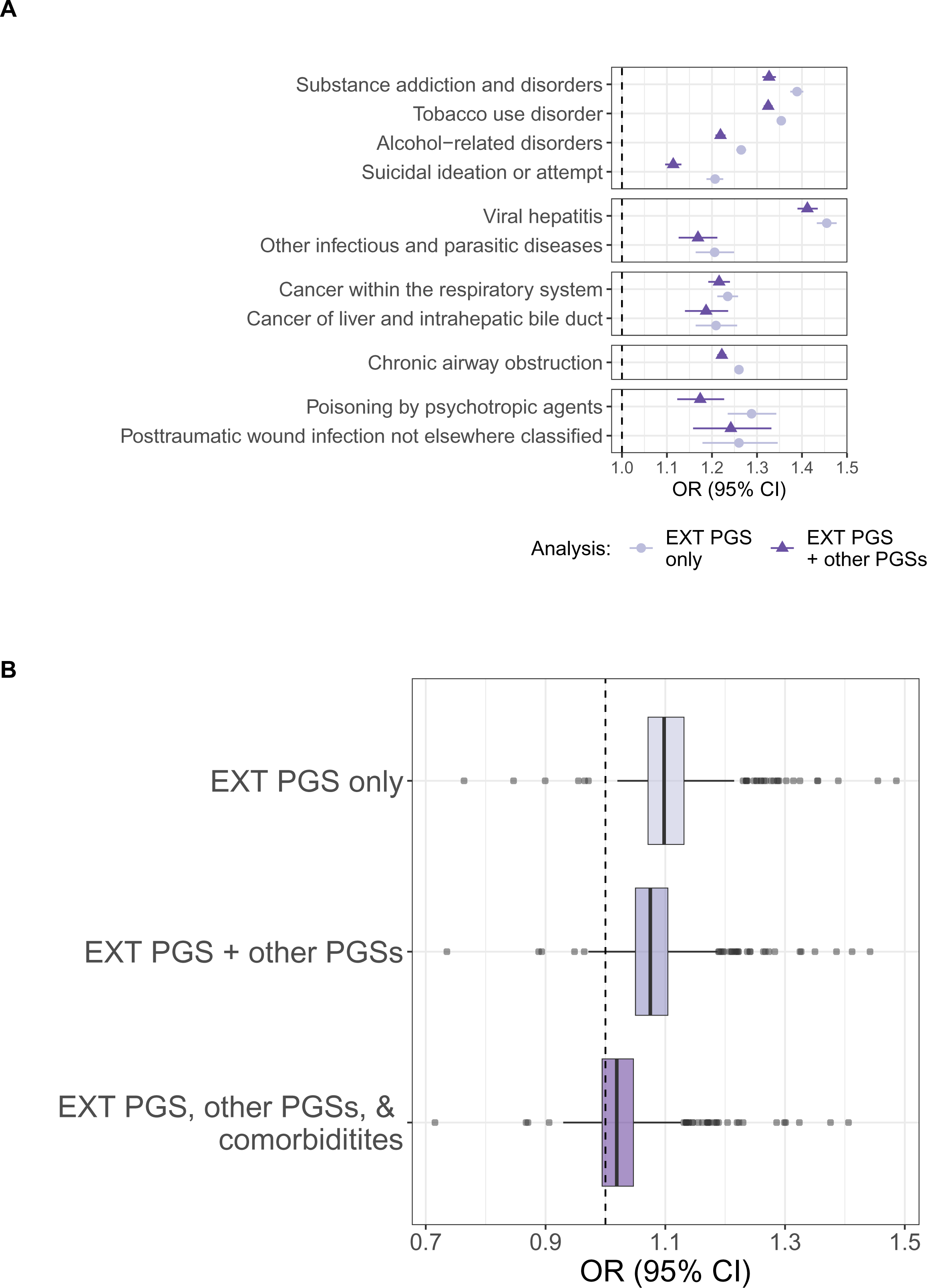
Associations between EXT PGS and Selected Phecodes Accounting for DEP, SCZ, and SUI PGS. Panel A presents selected associations and their corresponding effect sizes from conditional PheWAS (EXT + DEP, SCZ, and SUI PGS) in veterans of primarily European ancestries (N = 406, 254) compared to models with the EXT PGS only (marginal). Panel B presents box plots for the effect sizes from the 619 significant associations identified in the main PheWAS in: 1) the PheWAS of EXT PGS only; 2) the PheWAS of EXT and other PGSs; and 3) the PheWAS of EXT, other PGSs, and the total comorbidity score.

**Figure 4:**
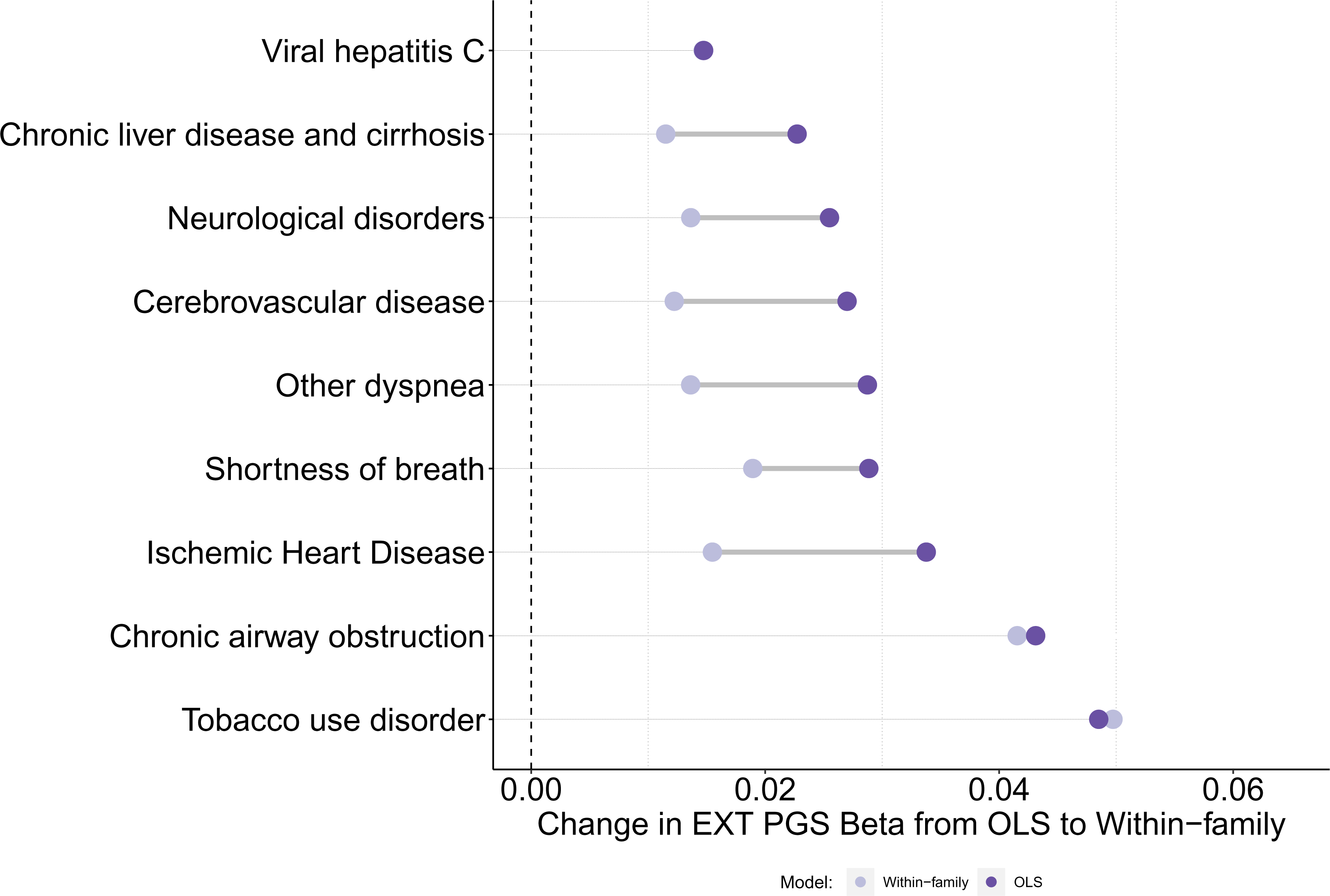
Change in Effect Sizes for EXT PGS in Within-family Models. Change in effect sizes for significant associations in a sample of related veterans of primarily European ancestries (*N* = 12,127). Estimates represent the change between ordinary least squares models (with clustered standard errors) and family fixed effects models. All associations significant after correcting for a false discovery rate (FDR) of 5%.

### Within-family replication of Main PheWAS Results

Finally, we investigated the 494 phecodes that remained significantly associated with EXT in the conditional PheWAS using a sample of full siblings from the broader MVP (*N* = 12,127). Of these 494 phecodes, 439 had available information in the subset of related veterans. Within these remaining 439 associations, 77 were marginally significant (*p* < .05) but only 13 of these associations remained after correcting for multiple testing using a false discovery rate (FDR) of 5%. Though many of the SUD phecodes were strongly associated in the main PheWAS and marginally significant in the within-family associations, only the tobacco use disorders association survived corrections for multiple testing (β*_within_* = 0.049, *p* < 1.28×10^-7^, see eTable 6 for full results).

Interestingly, the associations that remained significant within-family were overwhelmingly related to downstream consequences of various forms of substance use, including viral hepatitis C, chronic liver disease and cirrhosis, chronic airway obstruction, and ischemic heart disease. Figure 3 shows the relative effect sizes from the linear probability models with (ordinary least squares, or OLS) and without (within-family) family fixed-effects. For some of the associations we see a marked decrease in effect size. Therefore, a non-trivial portion of the association between the EXT PGS and these phenotypes may be due to some type of confounding. However, for viral hepatitis C, tobacco use disorder, and chronic airway obstruction, there is little or no attenuation, and these estimates could reflect some type of causal pathway between underlying risk and disease state.

## DISCUSSION

Problems related to behavioral disinhibition, commonly referred to as externalizing, may have detrimental consequences for health and well-being. We and others have shown that risk for EXT disorders overlaps with a variety of key public health outcomes at both the genetic and phenotypic level ^11,13–18,50^. In the current analysis, we leveraged these recent, novel insights into the underlying genetic basis of EXT, extended analyses to multiple ancestries, and evaluated their correlates in the largest integrated healthcare system in the US.

As expected, some of the strongest associations across all of the model specifications were in the phecode domain of *mental disorders*. Specifically, substance use disorders (alcohol, tobacco, and other substances) were among the strongest associations, though other psychiatric problems, including conduct disorder, personality disorders, and attention deficit/hyperactivity disorder (ADHD) were also associated. Some components, specifically problematic alcohol use, smoking, and ADHD, contributed to the multivariate GWAS used for creating the EXT PGS ^11^, but analyses in the original EXT paper showed that the latent factor was not driven by any single indicator, so it is unlikely that associations are driven any indicator-outcome similarity. Overall, these associations help to validate the EXT PGS within MVP and further demonstrate the utility of focusing on shared risk across psychiatric conditions^7^.

Results from the main PheWAS also replicated the diversity of bodily systems that were associated with risk for externalizing disorders ^11^. . For example, the EXT PGS remained associated with suicide-related phecodes (suicidal ideation, suicide attempt, and self-harm) even when conditioning on the suicide PGS and other comorbidities, suggesting that the association between externalizing and suicidal behaviors is independent of risk for the other forms of psychiatric problems (e.g., depression), supporting the role of impulsivity in suicide risk ^51^. In total, 181 (∼96%) of the associations that survived correction for multiple testing and covarying for other PGS and comorbidities were phenotypes traditionally used to measure externalizing (e.g., ADHD, SUDs). Overall, these results point to a robust pathway between risk for externalizing disorders and numerous medical conditions that: 1) replicated across ancestry, 2) was not explained by risk for other common forms of psychiatric problems, and 3) could not be fully explained by documented comorbidities.

The EXT PGS was associated with *reduced* risk for 6 phecodes: prostate cancer, melanoma, disorders of bilirubin excretion, autism, celiac disease, and flat foot. While associations may reflect a true reduced risk, the negative associations may also reflect our use of lifetime diagnosis. Because several of these diseases are strongly age-graded, supposedly protective associations may reflect bias due to mortality selection. Future work can leverage the longitudinal EHR data in MVP to characterize the association between and early mortality to determine whether there is any true protective effect.

The expansive MVP cohort allowed us the opportunity to explore the possibility of confounding influences via a *novel* approach: leveraging an appreciable number of full-siblings that comprise less than 2% of the overall cohort. The within-family analyses presented an additional test of whether EXT is simply a correlate or potentially causally related to various phecodes. In the holdout sample of full siblings, only a portion of the associations (13/439, 3.0%) remained associated after correcting for multiple testing. While suicide ideation, attempt and self-harm was one of the stronger associations in the PheWAS, it did not replicate on a within-family basis. This may have been due to the relatively few cases of suicidal ideation, attempt or self-harm in the EHR of the smaller within-family sample (n = 205, 1.7%). In terms of SUDs, only tobacco use disorders remained significant after multiple testing correction, though alcohol related disorders (phecode 317) was marginally significant. There was insufficient within-family variation to include substance and addiction disorders (phecode 316) in the within-family models. Moreover, many of the associations that replicated within-family were likely *consequences* of SUDs. These included chronic airway obstruction (smoking-related), chronic liver disease and cirrhosis (alcohol-related), and viral hepatitis C (intravenous drug use related). The within-family associations point to the potential causal impact of risk for externalizing on these medical conditions, likely mediated through SUDs. It is important to note that even though within-family associations are robust to confounding, they can be biased in the presence of genetic nurture and sibling effects ^52^.

It is important to note that even though we detected a large number of significant associations with the EXT polygenic score, many of the effect sizes are too small to be clinically relevant. The utility of polygenic scores in clinical settings is an ongoing discussion ^53,54^. Recent work in substance use disorders has shown that PGS are not sufficiently powered to meaningfully identify individuals at increased risk of developing SUDs^55^, especially when well-known social or clinical risk factors are included in the same model^50^.

Our analysis has several important limitations. First, although we included large samples of multiple ancestries, PGS were derived from a GWAS of primarily European ancestries. Consistent with recent observations of other PGS in MVP ^25^, the EXT PGS was associated with many of the traits within the AFR and HIS samples, but the effect sizes were highly attenuated. Large-scale discovery GWAS in diverse cohorts are vital to ensuring that PGS perform as well in these groups and that any benefit of precision medicine is shared equitably across the population ^56^. Second, results from this sample may not be generalizable to the broader U.S. population. While generally representative of the VA, MVP is still a selected subset comprising primarily male individuals. Additional work is needed to ensure that the study results generalize beyond the VA. Third, we did not examine PGS in conjunction with social and environmental factors. Both polygenic and environmental risk factors are important for understanding key outcomes in veterans’ health, including substance use disorders ^50^, major depressive disorder ^57^, and other mental health problems ^58^. Many veterans are at risk for adverse environmental experiences due to poverty, minority status, and physical and psychiatric challenges ^59^. Future work should endeavor to move beyond the focus on genetic or environmental risk in isolation and towards integrated approaches. Lastly, while our results are largely robust to additional confounds and replicate within-family, we cannot completely rule out non-causal reasons for the observed associations.

Risk for externalizing disorders is correlated to many outcomes of serious public health concern. A predisposition towards greater levels of externalizing is associated with increased risk of substance use disorders, suicide related behaviors, and other chronic medical conditions. Our analysis demonstrated that externalizing risk is equally important among the US veteran population who receive their healthcare within the VA system. Intervention and prevention efforts that identify ways to target and monitor the behavioral manifestations of externalizing risk (e.g., improving impulse control) could substantially improve morbidity and mortality outcomes.

## Supporting information

eTables

## Data Availability

Publicly available summary statistics are available through their respective consortia websites.

## Data Availability

Publicly available summary statistics are available through their respective consortia websites.

## ACKNOWLEDGEMENTS

The views expressed in this article are those of the authors, and do not necessarily reflect the position or policy of the Department of Veterans Affairs (VA). The Million Veteran Program (MVP) is funded by grant #MVP000 from the VA Office of Research and Development (ORD). This work was also funded by grant #1I01CX001729 from the Clinical Services Research & Development (CSRD) Service of VA ORD to Drs. Beckham and Kimbrel, by the MVP CHAMPION program, which is a collaboration between the VA and the Department of Energy (DoE), and by a CSRD Senior Research Scientist award (lK6BX003777) to Dr. Beckham. Drs. Barr, Bigdeli, Aslan, and Harvey are supported by VA Cooperative Studies Program (CSP) #572. Drs. Peterson, Bigdeli, and Meyers are supported by the National Institute of Mental Health (R01MH125938). Dr. Peterson is also supported by the National Institute on Alcohol Abuse and Alcoholism (P50AA022537) and the Brain Behavior Research Foundation NARSAD grant 28632 PS Fund. Drs. Barr and Dick are also supported by the National Institute of Drug Abuse (R01DA050721) and the National Institute of Alcohol Abuse and Alcoholism (R01AA015416). Dr. Sanchez-Roige was supported by funds from the California Tobacco-Related Disease Research Program (TRDRP; Grant Number T29KT0526 and T32IR5226) and the National Institute on Drug Abuse (DP1DA054394). Dr. Mallard is supported by funds from NIH T32HG010464. The content is solely the responsibility of the authors and does not necessarily represent the official views of the National Institutes of Health. Dr. Barr had full access to all the data in the study and takes responsibility for the integrity of the data and the accuracy of the data analysis.

We would also like to thank The Externalizing Consortium for sharing the GWAS summary statistics of externalizing. The Externalizing Consortium: Principal Investigators: Danielle M. Dick, Philipp Koellinger, K. Paige Harden, Abraham A. Palmer. Lead Analysts: Richard Karlsson Linnér, Travis T. Mallard, Peter B. Barr, Sandra Sanchez-Roige. Significant Contributors: Irwin D. Waldman. The Externalizing Consortium has been supported by the National Institute on Alcohol Abuse and Alcoholism (R01AA015416-administrative supplement), and the National Institute on Drug Abuse (R01DA050721). Additional funding for investigator effort has been provided by K02AA018755, U10AA008401, P50AA022537, as well as a European Research Council Consolidator Grant (647648 EdGe to Koellinger). The content is solely the responsibility of the authors and does not necessarily represent the official views of the above funding bodies. The Externalizing Consortium would like to thank the following groups for making the research possible: 23andMe Inc., Add Health, Vanderbilt University Medical Center’s BioVU, Collaborative Study on the Genetics of Alcoholism (COGA), the Psychiatric Genomics Consortium’s Substance Use Disorders working group, UK10K Consortium, UK Biobank, and Philadelphia Neurodevelopmental Cohort. We would like to thank the many studies that made these consortia possible, the researchers involved, and the participants in those studies, without whom this effort would not be possible. We would also like to thank the research participants and employees of 23andMe.

Most importantly, we would like to thank the U.S. Veteran participants in MVP for their service, and for their time, samples, and continued participation in VA research. Without them, this work would not be possible.

